# Evaluation of nasopharyngeal swab collection techniques for nucleic acid recovery and participant experience: recommendations for COVID-19 diagnostics

**DOI:** 10.1101/2020.08.18.20177592

**Authors:** Natalie N. Kinloch, Aniqa Shahid, Gordon Ritchie, Winnie Dong, Tanya Lawson, Julio S.G. Montaner, Marc G. Romney, Aleksandra Stefanovic, Nancy Matic, Chanson J. Brumme, Christopher F. Lowe, Zabrina L. Brumme, Victor Leung

**Affiliations:** Faculty of Health Sciences, Simon Fraser University, Burnaby, V5A 1S6, Canada; British Columbia Centre for Excellence in HIV/AIDS, Vancouver, V6Z 1Y6, Canada; Division of Medical Microbiology and Virology, St Paul’s Hospital, Vancouver, V6Z 1Y6, Canada; Department of Pathology and Laboratory Medicine, University of British Columbia, Vancouver, V6T 1Z7, Canada; Department of Medicine, University of British Columbia, Vancouver, V6T 1Z3, Canada

**Keywords:** COVID-19, nasopharyngeal swab, ddPCR, RT-ddPCR, participant experience, biological material, sample quality, ethnicity

## Abstract

Nasopharyngeal swabs are critical to the diagnosis of respiratory infections including COVID-19, but collection techniques vary. We compared two recommended nasopharyngeal swab collection techniques in adult volunteers and found that swab rotation following nasopharyngeal contact did not recover additional nucleic acid (as measured by human DNA/RNA copy number). Rotation was also less tolerable for participants. Notably, both discomfort and nucleic acid recovery were significantly higher in Asians, consistent with nasal anatomy differences. Our results suggest that it is unnecessary to rotate the swab in place following contact with the nasopharynx, and reveal that procedural discomfort levels can differ by ethnicity.

**summary:** Nasopharyngeal swabs are critical to COVID-19 diagnostics, but collection techniques vary. Comparison of two collection techniques revealed that swab rotation did not recover more nucleic acid and was more uncomfortable. Discomfort and biological material recovery also varied by participant ethnicity.

## Main text

Nasopharyngeal swabs are critical for accurate diagnosis of respiratory tract infections including COVID-19 [1, 2]. Specimen collection, which involves inserting a long flexible swab through the nostril along the floor of the nasal cavity to a depth of approximately 7 centimetres and into the nasopharynx, must be performed by a trained healthcare professional familiar with the technique and nasal anatomy [1]. There is however no consensus for optimal swab collection. Following contact with the nasopharynx, the WHO for example recommends that the swab be left in place for a few seconds before withdrawal [3] while the US-CDC recommends that the swab be “gently rubbed and rolled” and left in place for several seconds before withdrawal [4]. Other guidance documents recommend that the swab be rotated in place before removal [5-8]. Given that swab insertion/removal is invasive and uncomfortable [1, 9], a better understanding of the impact of post-insertion collection techniques on sample quality and patient experience may refine collection methods.

We recruited adult volunteers to undergo a nasopharyngeal swab with or without rotation, and to provide saliva, an alternative COVID-19 diagnostic specimen [10-12], as a comparator. Participants rated their discomfort during the swab on an 11-point scale [13] and were asked which specimen, swab or saliva, was less unpleasant to give. We assessed nucleic acid recovery as a marker of swab collection quality [2], where human RPP30 [2] and human RNase P copy numbers were used as surrogates for DNA and RNA recovery, respectively.

## Methods

We recruited 69 participants over three days in July 2020. A single, experienced healthcare provider collected all nasopharyngeal swabs using the Puritan® UniTranz-RT® transport system (Puritan Medical Products). Participants were alternately assigned to one of two swab collection techniques (see below), and were not informed of the technique until immediately prior to the procedure. Prior to collection, the provider instructed participants to alternatingly apply pressure to each nasal ala to identify the less congested nostril. The provider estimated the depth to the participant’s posterior nasopharynx by holding the swab externally from the nasal ala to tragus and viewed the nasal passage to check for mucus and obstructions (if mucus was visible the participant was instructed to blow their nose [1]). With the participant’s head tilted back slightly, the provider gently inserted the swab into the identified nostril along the lateral aspect of the nasal cavity floor and into the nasopharynx. For half the participants, the swab was removed after reaching the nasopharynx (“in-out” swab). For the remaining participants, the swab was rotated in place for 10 seconds following placement in the nasopharynx (“rotation” swab) and then removed. Swabs were immediately placed in transport medium. Participants were also asked to provide approximately 2mL saliva into a sterile container (Starplex® Scientific) by focusing on pooling saliva and gently expelling it into the container, repeating until the required volume was achieved. Participants were asked to rate their discomfort during the swab on an 11-point scale [13], where “0” denoted a complete absence of discomfort and “10” denoted the most severe discomfort possible. Finally, participants were offered the hypothetical choice of providing saliva or undergoing a nasopharyngeal swab for a diagnostic purpose, and asked: *purely based on your experience today*, which sample would you prefer to give and why?

Swabs were processed within 5 hours of collection. Total nucleic acids were extracted from 1mL of medium on a NucliSens easyMAG (BioMérieux) and eluted in 60μL. Eluates were split into three aliquots and frozen at −80°C until use. Droplet digital PCR (ddPCR) and Reverse-Transcriptase (RT)-ddPCR were used to quantify human RPP30 copy numbers and RNase P transcript numbers, respectively. In this technology, each sample is fractionated into 20,000 nanolitre-sized water-in-oil droplets prior to amplification with sequence-specific primers and fluorescent probes, and input template concentrations are calculated at endpoint using Poisson statistics. The RPP30 assay, described previously, yields a final measurement of cells/μL extract [2]. In the RNase P assay, nucleic acid extracts were combined with the US-CDC-developed RNAseP-specific primer/probe set [14], XhoI restriction enzyme (New England Biolabs) and the One-Step RT-ddPCR Advanced Kit for Probes (BioRad). Primer and probe sequences are as follows: Forward Primer-AGATTTGGACCTGCGAGCG, Reverse Primer-GAGCGGCTGTCTCCACAAGT, Probe-FAM-TTCTGACCT-ZEN-GAAGGCTCTGCGCG-3IABkFQ (Integrated DNA Technologies; ZEN=internal ZEN quencher; 3IABkFQ=3’ Iowa Black Quencher). Droplets were generated using an Automated Droplet Generator (BioRad) and cycled at 50°C for 60 minutes; 40 cycles of (94°C for 30 seconds, 55°C for 1 minute) and 98°C for 10 minutes and analyzed on a QX200 Droplet Reader using QuantaSoft software version 1.7.4 (BioRad). Measured RNase P copies were normalized to input volume to determine RNase P copies/μL extract. RNase P levels were also assessed using real time reverse transcriptase (RT)-PCR with the same primer/probe set on a Roche Lightcycler 480 according to the US-CDC protocol [15]. All three assays were performed on independent extract aliquots to avoid freeze-thaw. All ddPCR and RT-ddPCR assays were performed in duplicate, and results averaged between replicates. Non-parametric statistics were used for all correlations and between-group comparisons. Contingency tables were analysed using Fisher’s exact test. Lin’s concordance coefficient was used to calculate concordance between replicates.

This study was approved by the Providence Health Care/University of British Columbia and Simon Fraser University Research Ethics Boards. All participants provided written informed consent.

## Results

The median age of the 69 participants was 42 years (Interquartile range [IQR] 36-54 years); 43 (62%) were female. Self-reported ethnicities were 44 (64%) White, 21 (30%) Asian (including 14 East, 1 Central, 3 South and 3 South-East), and 4 (6%) Other (including 2 Latino and 2 mixed-ethnicity). For 5 (7.2%) participants, the nasopharyngeal swab was unsuccessful due to obstruction despite attempts through both nares, leaving 64 (34 “in-out” and 30 “rotation”) swabs for analysis. Discomfort scores ranged from 1 (minimal discomfort) to 10 (maximum discomfort) in both swab groups, with no significant difference between them (median 5 [IQR 3.75-5] for “in-out” versus 4.5 [IQR 4- 6] for “rotation”, p=0.51) (Figure 1A). This suggests that most of the discomfort occurs during swab insertion/withdrawal, a notion that is supported by the significantly higher discomfort reported by participants with occlusions (p<0.001; Figure 1B). However, responses to additional study questions suggested that swab rotation was less tolerable. Firstly, though most participants preferred giving saliva, 10 of 34 (29.4%) participants in the “in-out” group preferred the swab, versus only 3 of 30 (10%) of participants in the “rotation” group (Fisher’s exact test p=0.068; Figure 1C), citing that the swab was easier, faster and/or generally less unpleasant than giving saliva. Moreover, two participants in the “rotation” group mentioned that they had previously undergone an “in-out” swab, and that rotation was more uncomfortable. One added that, given the choice between “in-out” swab and saliva, that they preferred the swab, but given the choice between “rotation” swab and saliva, they preferred saliva. Discomfort scores did not differ by sex (p=0.85) or age (Spearman’s ρ= 0.05, p=0.7). Of note however, Asian participants reported significantly higher discomfort scores compared to White participants (median 5 [IQR 4-7] versus 4 [IQR 3.5- 5] respectively p=0.047, Figure 1D).

**Figure 1:**
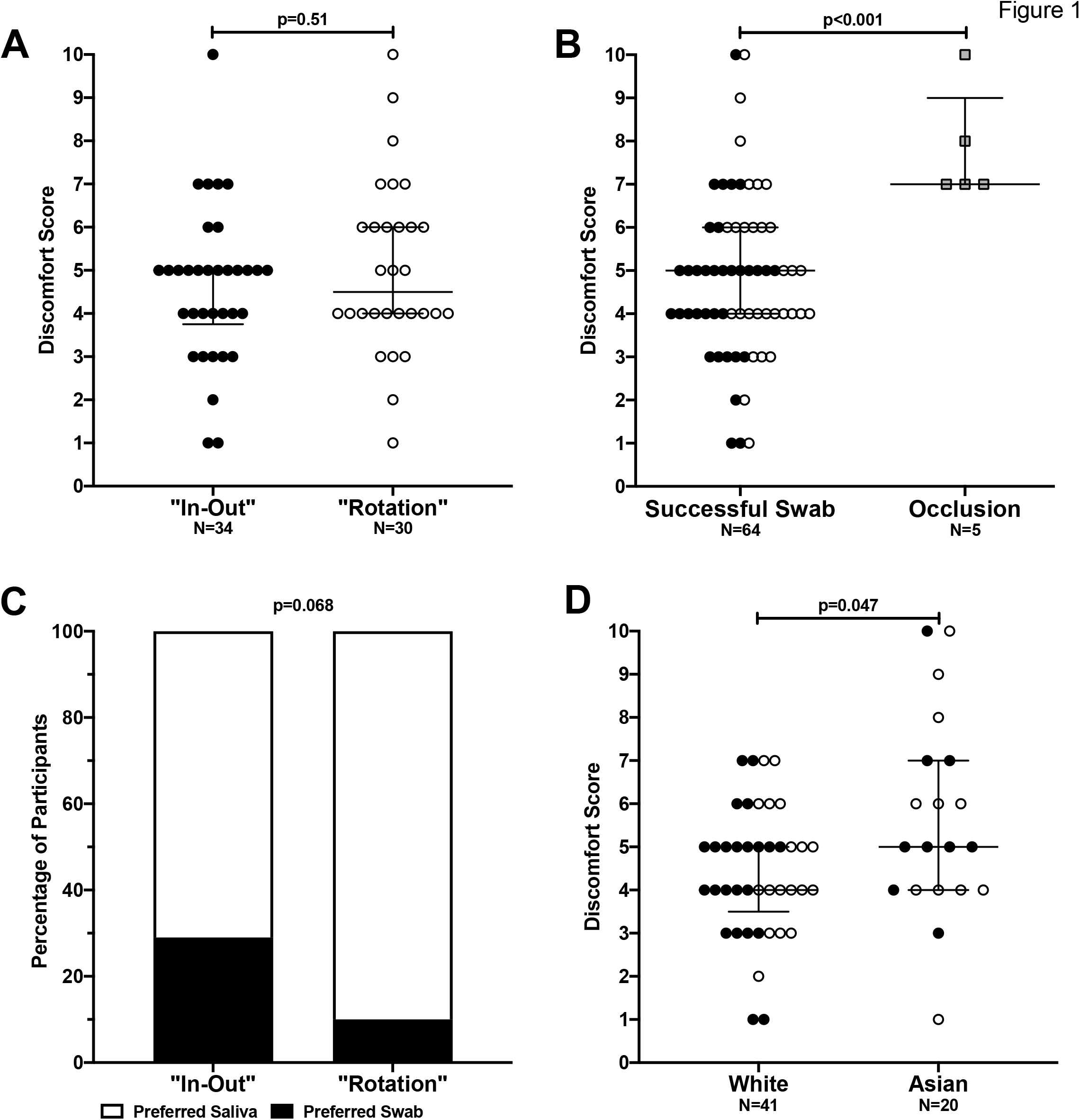
Differences in participant experience by nasopharyngeal swab technique and ethnicity. **(1A)**. No difference in discomfort score between “in-out” (black circles) and “rotation” (white circles) swab groups was observed. **(1B)**. Significantly higher discomfort was reported in participants with occlusion (grey squares) compared to those with a successful swab (black and white circles, denoting the groups described in 1A). **(1C)**. A greater proportion of “rotation” swab participants preferred to give saliva compared to “in-out” swab participants. **(1D)**. Significantly higher discomfort scores were reported in Asian compared to White participants. Individuals of other ethnicities (Latino, mixed ethnicity) were excluded due to low numbers (n=4).

RPP30 (DNA) and RNase P (RNA) copy numbers were measured as surrogates of nucleic acid recovery. Concordance between replicates was high for both targets (Lin’s Concordance Coefficient: RPP30=0.98, p<0.0001; RNase P=0.91, p<0.0001) and measurements of both targets correlated strongly with one another (Spearman’s ρ=0.84, p<0.0001). Both RPP30 and RNase P levels varied markedly regardless of swab technique: RPP30 levels extended over a 42-fold range (from 42 to 1751 cells/μL extract; Figure 2A) while RNase P levels extended over a 20-fold range (from 183 to 3570 copies/μL extract; Figure 2B). Moreover, we found no significant differences in RPP30 levels by swab technique (median 500 [IQR 235- 738] cells/μL extract for “in-out” versus 503 [IQR 398- 685] for “rotation”, p=0.83; Figure 2A), or RNase P values by swab technique (median 1338 [IQR 610- 2039] RNase P copies/μL extract for “in-out” versus 1309 [IQR 973- 1789] for “rotation”, p=0.84; Figure 2B). Together, this indicates that swab rotation does not recover more nucleic acid, and suggests that the amount of cellular material recovered is participant-specific. Indeed, when stratified by ethnicity, nucleic acid recovery was significantly higher in Asians: median RPP30 levels were 610 [IQR 430- 780] versus 431 [IQR 223- 621] cells/μL extract from Asian versus White participants (p=0.026; Figure 2C) while median RNase P levels were 1629 [IQR 1167- 2095] versus 1193 (531- 1758) RNase P copies/μL extract from these same groups (p=0.038; Figure 2D). No significant differences in RPP30 or RNase P levels were observed by sex, age or recruitment date (a surrogate of nucleic acid extraction run). RNAse P levels were also measured using the US-CDC 2019-nCoV real-time RT-PCR diagnostic assay [15]. The resulting cycle threshold (Ct) values correlated strongly with those measured using RT-ddPCR (Spearman’s ρ=-0.9, p<0.0001) and yielded results entirely consistent with those described above (data not shown).

**Figure 2:**
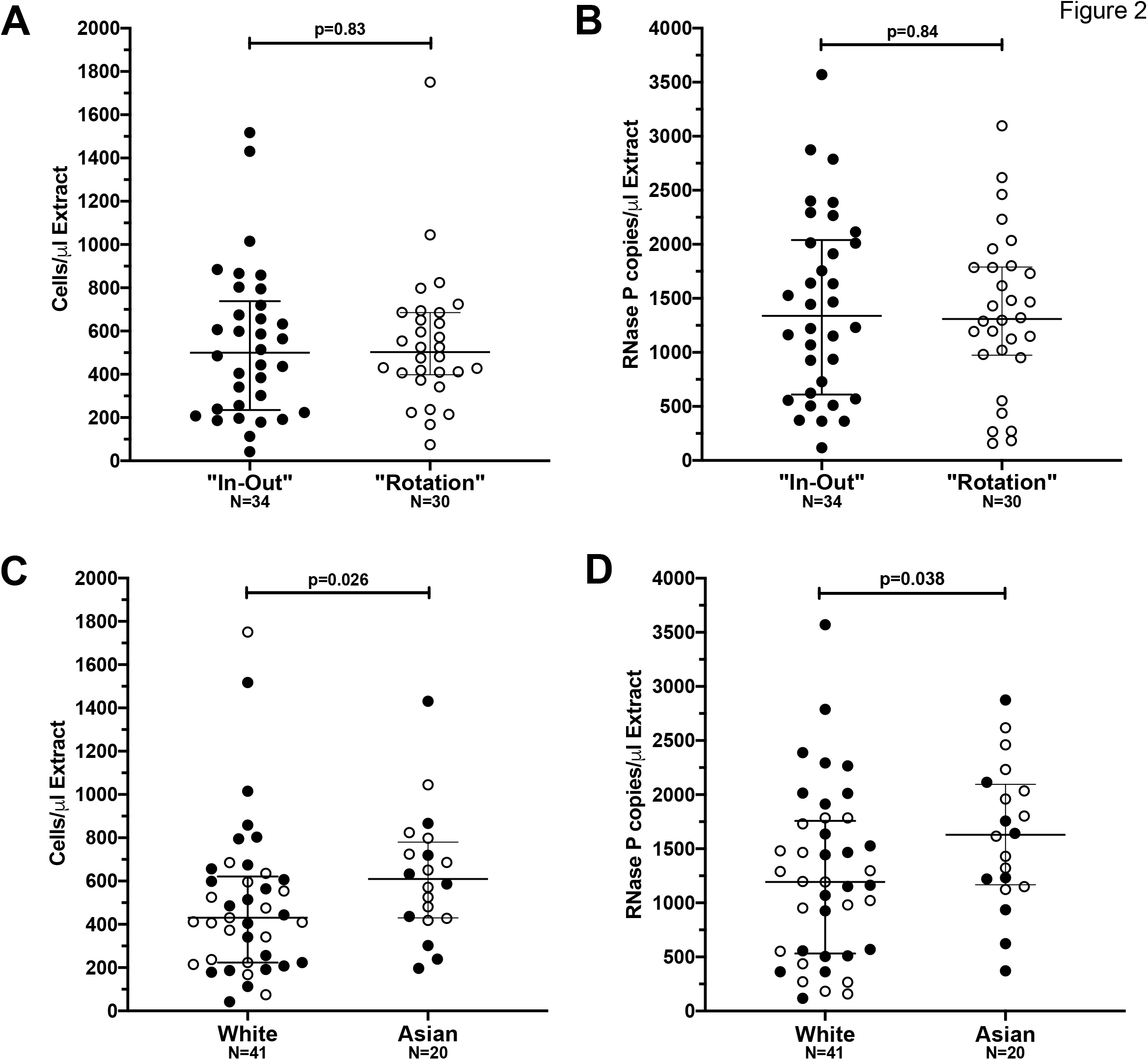
Differences in nucleic acid recovery by nasopharyngeal swab technique and ethnicity. **(2AB)**. No difference in DNA (RPP30, cells/μL extract, **2A**) or RNA recovery (RNase P, RNase P copies/μL extract, **2B**) between “in-out” (black circles) and “rotation” (white circles) swab technique groups was observed. **(2CD)**. Significantly higher levels of DNA (RPP30, cells/μL extract, **2C**) and RNA (RNase P, RNase P copies/μL extract, **2D**) were recovered on swabs from Asian compared to White participants. Individuals of other ethnicities (Latino, mixed ethnicity) were excluded due to low numbers (n=4).

## Discussion

Our observations have the potential to improve nasopharyngeal sample collection. For 7.2% of individuals, nasopharyngeal sampling was not possible due to obstructions (*e.g*. nasal polyps or deviated septum) and the procedure caused significantly more discomfort in these individuals. Providers should be aware of the frequency and discomfort implications of such occlusions, and should be issued appropriate guidance (*e.g*. do not force swab; sample ‘midturbinate’ area of the nasal cavity if the nasopharynx cannot be reached and note the swab location). Our observations further indicated that, despite being widely recommended, swab rotation upon contact with the nasopharynx does not enhance nucleic acid recovery and is less tolerable. We speculate that swab saturation is essentially achieved during entry and very brief resting in the nasopharynx, such that rotation does not recover additional material. Since we followed guidance to remove excess mucus prior to swabbing (as it can reduce the collection of desired cellular material [16]), our results are unlikely attributable to mucus collection.

The marked spread in discomfort scores was also notable. Though the average score in our study (5 on an 11-point scale) is similar to previous reports for nasopharyngeal swabs (an average of 3 on a 6-point scale [9]), the variation in discomfort from minimal to extreme underscores the need for providers to be mindful of inter-individual differences in experience. Intriguingly, Asian participants reported on average 1-point greater discomfort than White participants. This may be related to differences in the shape, contour and/or size of the nasal cavities and nasopharynx. Indeed, after adjustment for weight, age and sex, a study of facial anthropometric differences by ethnicity reported significantly lower nasal volumes (measured at 0-4cm from nostril), lower mean cross-sectional nasal area (at 0-6cm), and longer distances to the minimal cross-sectional area in Asian compared to White individuals [17]. Enhanced nucleic acid recovery on swabs from Asian participants is also consistent with narrower nasal passages in Asian compared to White individuals [17] that could increase both swab discomfort and mucosal contact, though marked variation in nucleic acid recovery across all ethnicities was noted. Nevertheless, healthcare providers should be sensitive to differences in discomfort levels across diverse populations.

Some limitations of our study merit mention. We assume that human DNA/RNA targets are appropriate markers of respiratory pathogen collection quality [2], though this is consistent with the inclusion of RNase P in the US-CDC 2019-nCoV real-time RT-PCR diagnostic panel as part of quality control [15]. As swabs vary in design, the absolute discomfort scores and nucleic acid quantities recovered may not be applicable to all swabs, though our general observations should be. Protocol differences also prevent direct comparison of recovered nucleic acid across studies (*e.g*. the swab, silica input and elution volumes differed between the present and a previous study by our group [2]).

## Conclusion

When performing nasopharyngeal sampling, rotation of the swab upon contact with the nasopharynx does not enhance sample quality. The observation that procedural discomfort levels differ significantly by ethnicity underscores the need for care providers to be sensitive to such differences, and more broadly underscores the importance of diverse participant representation in health research. Review and standardization of nasopharyngeal swab collection guidance notes should be a priority in the current COVID-19 pandemic.

## Data Availability

All data described in this manuscript are available upon reasonable request to the corresponding author.

## Acknowledgements

We thank the laboratory teams at the St. Paul’s Hospital Virology Laboratory and the BC Centre for Excellence in HIV/AIDS and Dr. Wayne Vogl for technical assistance.

